# Evidence-based interventions targeting mental health problems in adolescents living with HIV: A scoping review

**DOI:** 10.1101/2025.03.01.25323162

**Authors:** Van Thi Ngoc Tran, Lam Khanh Phung, Hoa Hong Nguyen, Linh Thi Dan Pham, Diep Thi Ngoc Nguyen, Thao Thi Thu Nguyen, Van Thi Hai Hoang, Bradley Neil Gaynes

## Abstract

**Introduction:** Adolescents living with HIV are at an increased risk of experiencing mental health challenges, which may impact their overall well-being and adherence to treatment. Evidence-based interventions are crucial to addressing these issues; however, the effectiveness of these interventions remains unclear. This scoping review aimed to synthesize all interventions that tested either the prevention or improvement of mental health for adolescents living with HIV.

**Methods:** We used PubMed, PsycINFO, CINAHL, Embase, and Cochrane to identify RCTs evaluating mental health interventions for HIV-infected adolescents.

**Results:** A scoping review included 13 out of 1015 studies demonstrating the global relevance of addressing mental health in this population. Interventions were diverse and showed mixed effectiveness in improving mental health outcomes such as depression, anxiety, trauma, and behavioural symptoms. Factors contributing to mixed results included variations in intervention design, study characteristics, and contextual factors. Identified gaps in the literature encompassed the limited number of studies in some regions and the lack of research on specific subpopulations and long-term intervention effectiveness.

**Conclusion:** A mixed result needs to be confirmed in future RCTs. This review provides valuable insights into improving the mental health of HIV-infected adolescents and can guide further research and practice in this area.

## Introduction

Mental health is a crucial component of overall well-being, and its importance is especially pronounced for adolescents living with HIV. In comparison to the general population, people living with HIV have a higher prevalence of mental health issues such as depression and anxiety [1]. Unique challenges faced by HIV-infected adolescents, such as stigma, discrimination, and difficulties adhering to antiretroviral therapy (ART), contribute to these increased mental health concerns [2, 3]. To address these issues and support the well-being of HIV-infected adolescents, it is imperative to identify and implement effective, evidence-based interventions targeting mental health outcomes in this population.

Despite the pressing need for such interventions, a notable gap exists in the literature concerning their effectiveness specifically tailored for adolescents living with HIV. Previous reviews have primarily focused on adult populations or mixed adolescents and adults who are either HIV-infected or HIV-affected [4, 5]. As a result, determining the effectiveness of these interventions for HIV-infected adolescents remains challenging. Given the unique psychosocial contexts and critical developmental stages experienced by adolescents, it is crucial to evaluate the effectiveness of interventions designed for this population.

This scoping review aimed to provide an evidence-informed guide for future research and practice by identifying the evidence base for mental health interventions in HIV-infected adolescents (10-19 years) and providing a summary of their characteristics and effectiveness. Specifically, we evaluated the current knowledge relating to the effectiveness of mental health interventions for adolescents living with HIV. The results will elucidate the mental health challenges faced by HIV-infected adolescents and inform the suitable model interventions worldwide for these people.

## Materials and methods

The protocol for this review was registered in the PROSPERO with the registration ID CRD42022382740 [6]. We followed the PRISMA checklist for our scoping review [7].

### Search strategy

We searched for potential articles on 24^th^ December 2023 and updated them on 17^th^ September 2024, using five databases (PubMed, Embase, Cochrane, CINAHL, and PsycINFO). The major keywords for searching were the targeted population (adolescents, children, teenagers, youth, juvenile, HIV, AIDS), intervention (evidence-based intervention, evidence-based practice, evidence-based policy, implementation, strategy, program, project), outcome (mental health, mental disorders, depression, anxiety, behavior problem, mental problem, psychiatric illness, psychiatric disorders), and study design (controlled clinical trial, comparative study, implementation science).

### Inclusion and exclusion criteria

We focused on studies involving HIV-infected individuals aged 10-19 years where the intervention targeted mental health outcomes. We required a comparison group for each study; however, we did not require that studies involve patients with elevated mental health measures at baseline. We excluded those studies solely evaluating well-being outcomes (e.g., self-worth, self-confidence, self-esteem, stigma, quality of life). Clinical and comparative studies were included, while observational studies like cross-sectional studies were excluded. We considered full-text articles but not reviews, editorial letters, conference papers, abstracts, and case studies.

### Screening process

Two authors screened potential articles in December 2023. Two researchers updated screening articles in September 2024. All reviewers independently screened studies based on the same inclusion and exclusion criteria, with any discrepancies resolved through discussion and consensus. The screening process began with titles and abstracts and then the screening full text was done. We excluded articles that did not report mental health outcomes, inappropriate intervention (not aiming to solve a mental health problem), non-HIV-infected adolescents, and unavailable full text. All screening results were shared with all members of the research team.

### Data collection and statistical analysis

Data were extracted from each eligible study, including study characteristics (author, publication year, design, studied sample, comparison, follow-up duration, analytical techniques, and trial registration), participant demographics (mean age, female proportion, and baseline mental health metrics), intervention details, outcome measures, and study outcomes (efficacy of intervention relative to the comparator). A narrative synthesis was conducted to offer a comprehensive overview of existing evidence-based interventions that aimed at mental health outcomes within the defined population, with a specific emphasis on the characteristics of these interventions and their associated findings.

## Results

### Literature search

The scoping review identified 13 out of 1015 studies that met the inclusion criteria after screening and eligibility evaluation [8–20] (**Figure 1**).

**Figure 1:**
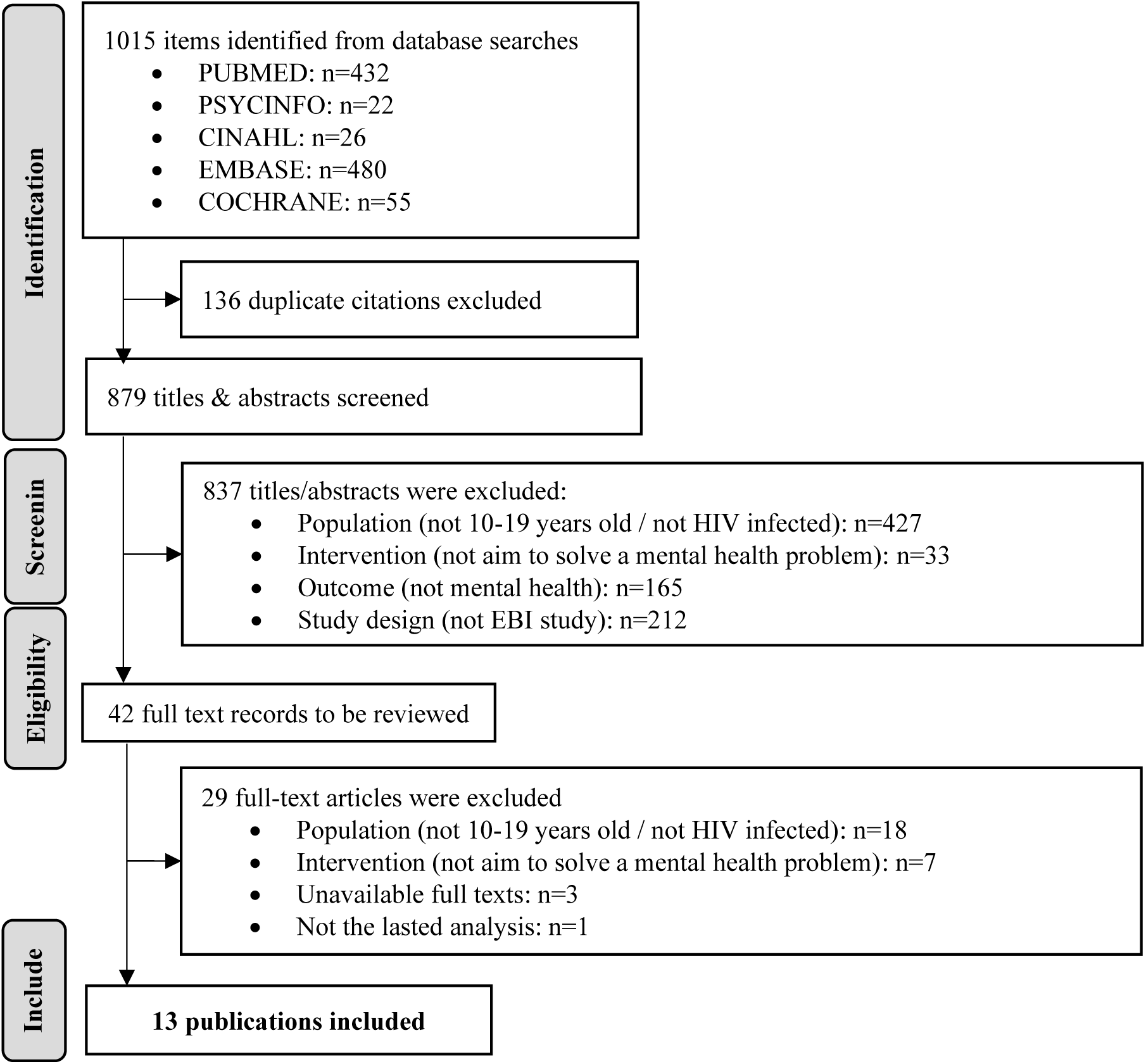
Prisma flowchart of the included studies.

### Study characteristics

**Table 1** highlights the characteristics of 13 studies examining mental health interventions for adolescents living with HIV. The majority (10/13) were published between 2019 and 2024, indicating a growing interest in targeted interventions for this population. These randomized controlled clinical trials (RCTs) had diverse sample sizes (21-842 participants) and included participants from countries like Botswana, Tanzania, Rwanda, Zimbabwe, Uganda, Kenya, Thailand, and the United States. Ages ranged from 15 to 19 years, and female representation varied with a maximum of 66%. Evaluated interventions were assessed using various scales. Follow-up periods ranged from 1 to 48 months, and analysis methods featured intention-to-treat (ITT) approaches, post-hoc models, mixed-effects models, and regression models, offering insight into the interventions’ impact on mental health outcomes over time and identifying factors associated with treatment response.

**Table 1.**
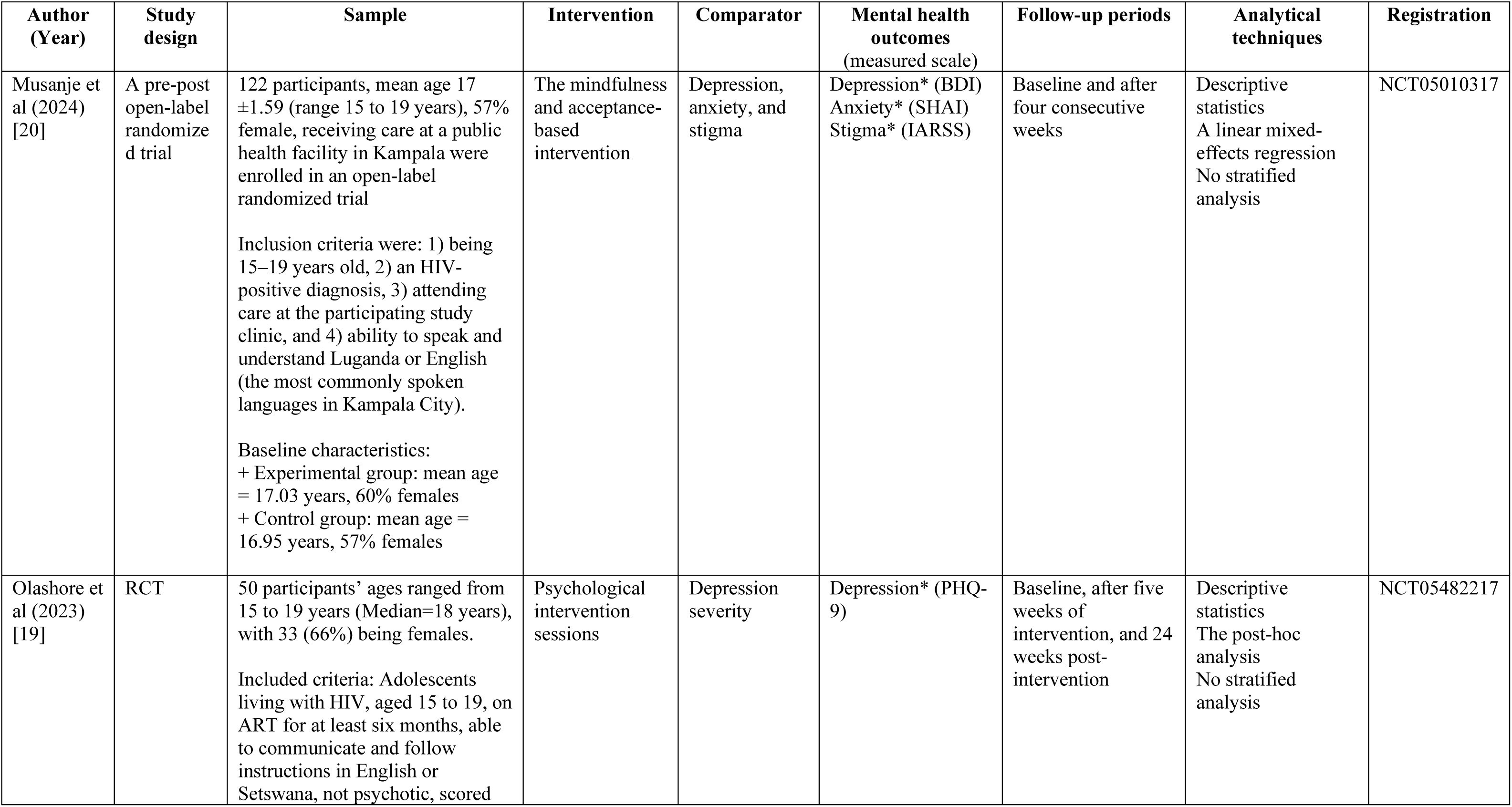
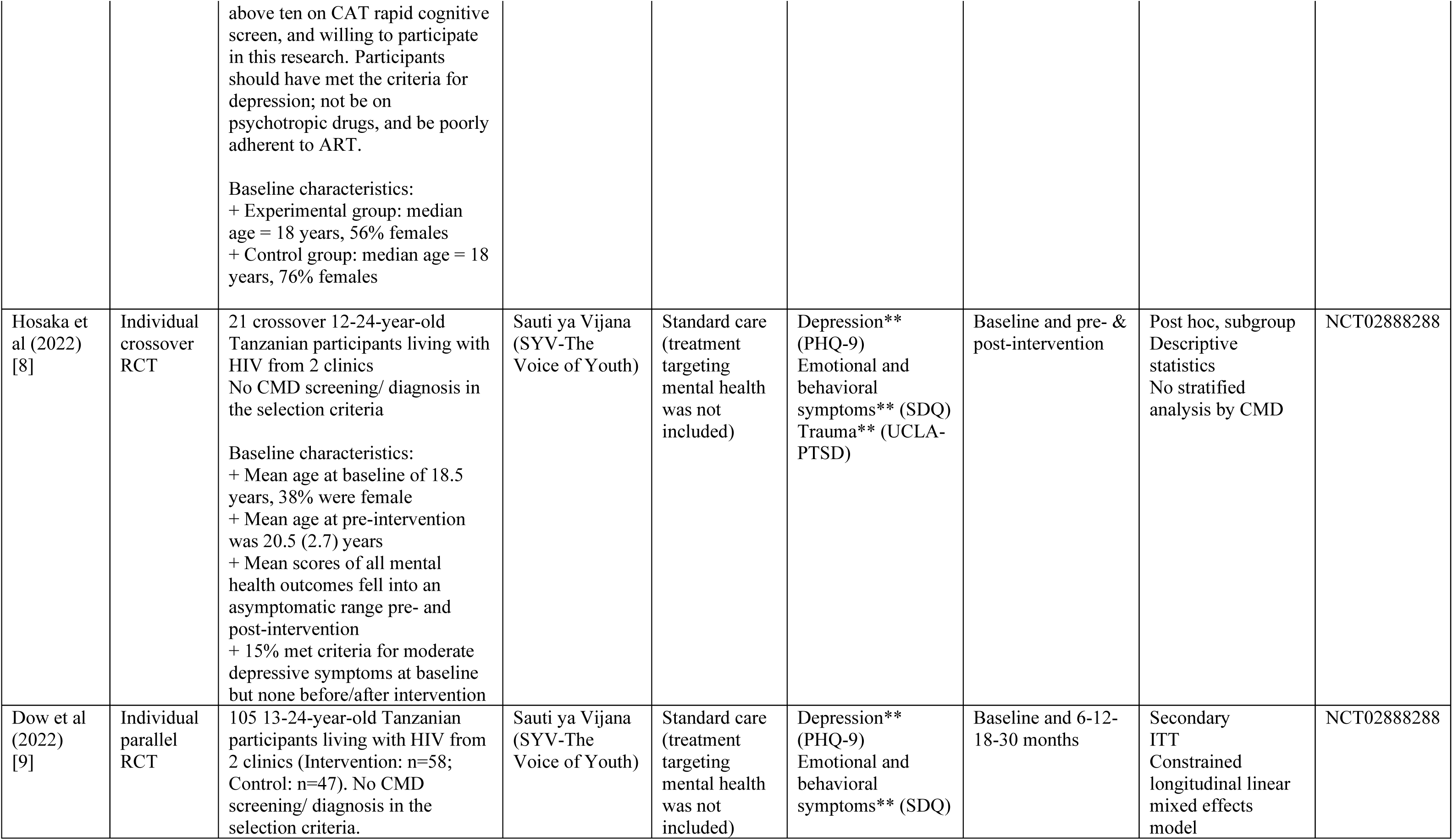
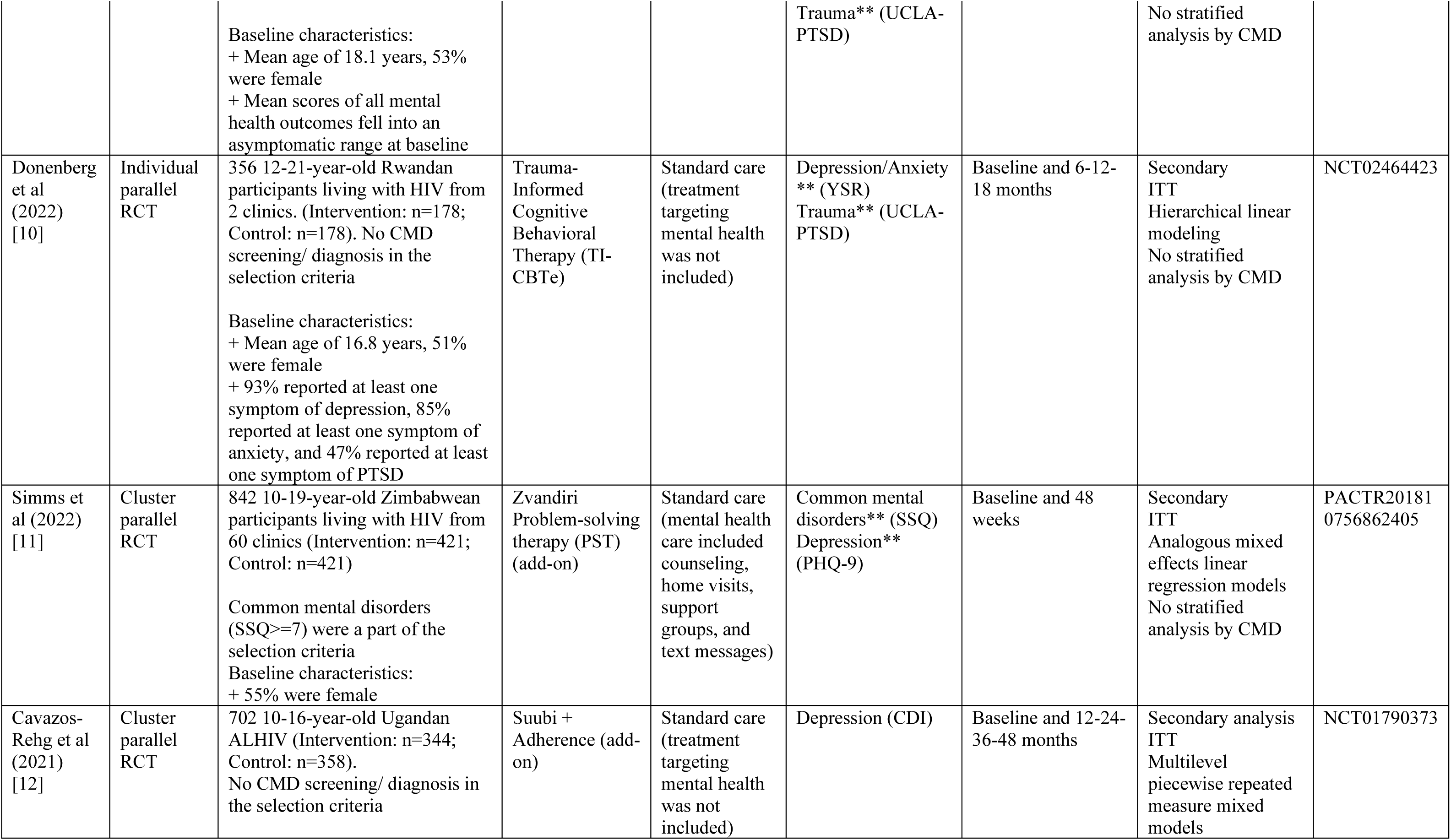
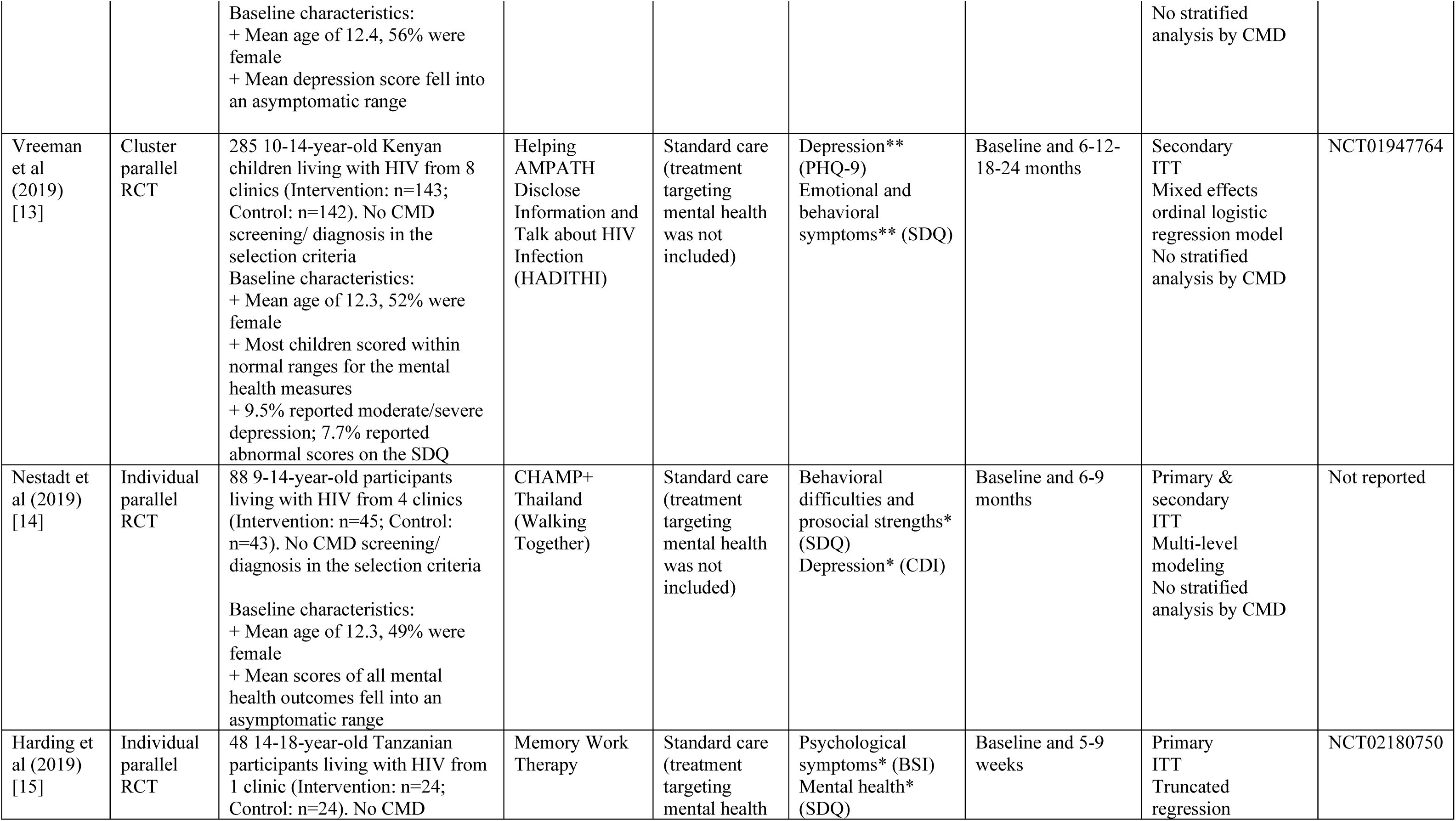
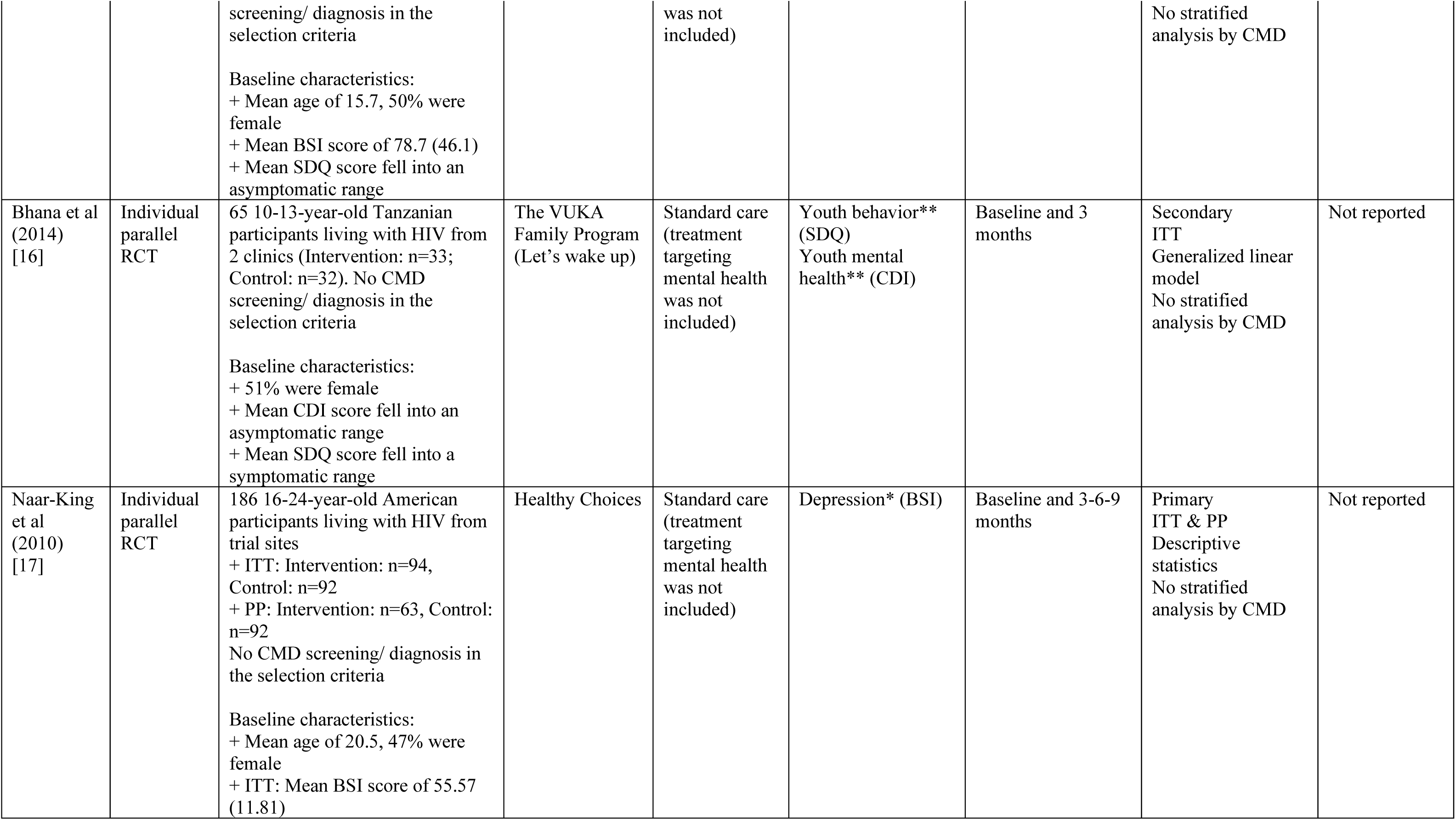
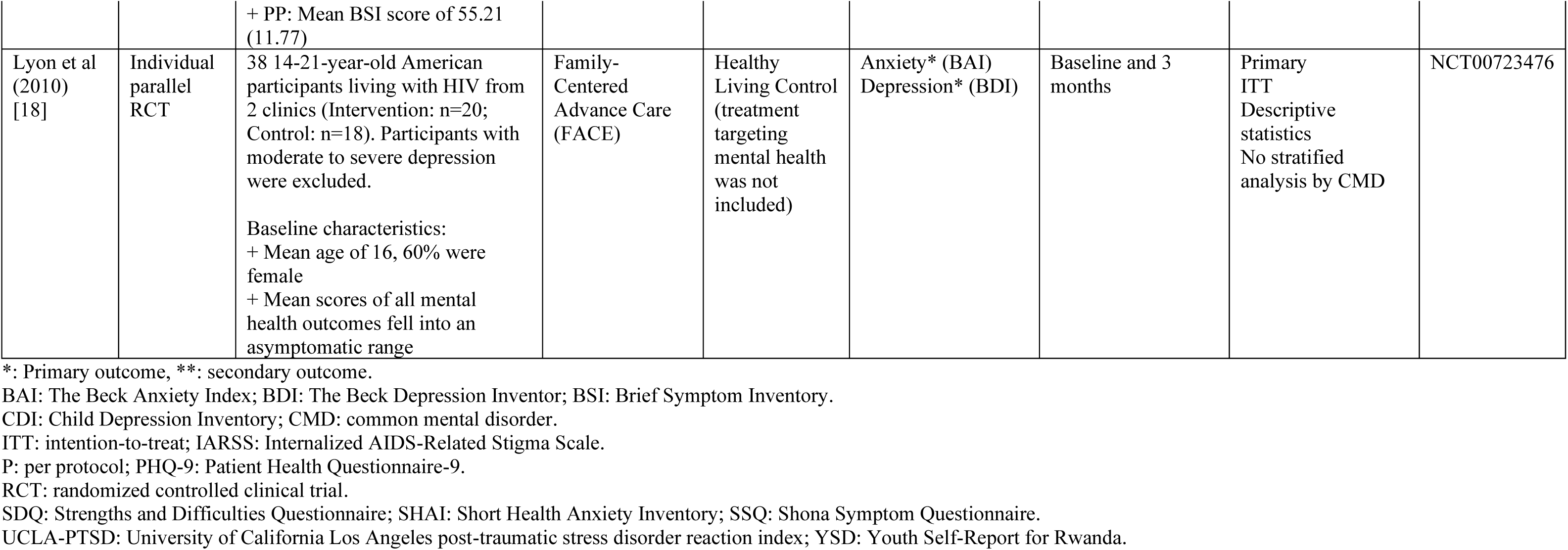
Characteristics of included studies (n=13)

### Characteristics of evidence-based interventions

**Table 2** describes the characteristics of ten interventions used in eleven studies addressing the mental health of adolescents living with HIV. These interventions varied in delivery mode, duration, provider, and treatment approach, typically employing psychotherapy/ behavioral or family-based and support approaches (involving caregivers). While some interventions were novel or based on pilot work, others integrated evidence-based approaches from the literature. Most interventions consisted of group and/or individual sessions, delivered weekly for several weeks to months by trained professionals or lay counselors with relevant experience. Types of interventions included mindfulness and acceptance-based intervention, psychoeducation intervention, cognitive behavioral therapy, trauma-informed therapy, problem-solving therapy, financial management, disclosure, advance care planning, and substance use/sexual risk/medication adherence.

**Table 2.** Characteristics of evidence-based interventions (n=12)

| Author (Year) | Intervention description | Delivery mode | Duration | Provider | Treatment approach | Participant components | Adaptation |
| --- | --- | --- | --- | --- | --- | --- | --- |
| Musanje et al (2024) [20] | The intervention group involved sessions of clarifying values, skillfully relating to thoughts, allowing and becoming aware of experiences non-judgmentally, and exploring life through trial and error. Additionally, this group also received the current standard of care. Meanwhile, the control group received the current standard of care only. | 90-minute group sessions for four consecutive weeks | Pre-intervention and after 4 weeks | Two experienced trainers | The mindfulness and acceptance-based intervention | Without caregivers | Four sessions of the intervention rather than six, recommended by local health experts |
| Olashore et al (2023) [19] | The intervention involved interactive discussion, roleplay, and brief plenary sessions in the local language, Setswana, and mixed with English. It included five sessions: 1) introduction to the program and rapport building, 2) identifying and dealing with good adherence I and II, 3) handling privacy, 4) social support, and 5) handling slips and wrapping up the review. | 60 minutes for each session (Five sessions). | Pre-intervention, 5 -and 24 weeks post-intervention | The team psychologist and the principal investigator | Psychoeducation and problem-solving | Without caregivers | Integration of intervention programs designed in high-income countries |
| Hosaka et al (2022) [8] & Dow et al (2022) [9] | Sauti ya Vijana (SYV-The Voice of Youth): 10 90-minute group sessions (2 with caregivers) and 2 individual sessions led by trained professionals using evidence-based treatment models like cognitive behavioral therapy (CBT), interpersonal psychotherapy (IPT), and motivational interviewing (MI) | 1090-minute group and 2 individual sessions | Weekly for 10 weeks | Trained group leaders who had HIV/mental health research experience | Psychotherapy and behavioral | With caregivers | Integration of several evidence-based models |
| Donenberg et al (2022) [10] | Trauma-Informed Cognitive Behavioral Therapy (TI-CBT): 6 2-hour group sessions including psychosocial health education, relaxation training, cognitive restructuring, adherence barriers, and caregiver psychoeducation | 62-hour group and individual sessions | Weekly for 8-10 weeks | Trained young Rwandans living with HIV | Psychotherapy and behavioral | With caregivers | Integration of several theories & approaches Steppingstones |
| Simms et al (2022) [11] | Zvandiri Problem-solving therapy (PST): 4 steps outlined in the Friendship Bench manual including (1) "kuvhura pfungwa" (open mind), client makes a list of problems; (2) "kusimudzira" (uplift), counselor helps choose relevant problem & brainstorm solutions; (3) "kusimbisa" (strengthen), select solution & create | Group and individual sessions | Every other week for 48 weeks | Zvandiri mentor | Family-based and support | With caregivers | Friendship Bench model |
|  | SMART action plan; (4) "kusimbisisa" (further strengthen), join support group |  |  |  |  |  |  |
| Cavazos-Rehg et al (2021) [12] | Suubi + Adherence: 4 sessions on financial management and training in income-generating activities and combined a matched savings account—opened in the child and caregiver names; and 12 educational sessions with a paired mentor, including, but not limited to, financial planning, business development, saving, setting short- and long-term goals, and avoiding risk-taking behaviors | Group sessions | 24 months | Lay counselors and finance mentor | Family-based and support | With caregivers | Integration of evidence-based approaches presented in the literature |
| Vreeman et al (2019) [13] | Helping AMPATH Disclose Information and Talk about HIV Infection (HADITHI): Disclosure intervention comprises access to intensive group and one-on-one counseling | Group and individual sessions | Not reported | Trained counsellors | Family-based and support | With caregivers | Previous pilot work |
| Nestadt et al (2019) [14] | CHAMP+ Thailand (Walking Together): 11 sessions using structured manualized intervention, including a cartoon-based curriculum, follow-up activities, and facilitated discussion | Group and individual sessions | Weekly for 6 months | Trained social worker or counselor | Family-based and support | With caregivers | CHAMP model |
| Harding et al (2019) [15] | Memory Work Therapy: A residential camp, including a Memory box, Memory book, Tree of life and the Hero (Active Citizen) book, to create a safe space for the participants to explore their life story | Group and individual sessions | 5 days | 5 staff (3 qualified social workers, 1 clinical officer, and 1 medical doctor) | Psychotherapy and behavioral | Without caregivers | Novel |
| Bhana et al (2014) [16] | The VUKA Family Program (Let's wake up): 10 sessions on a cartoon-based story of a 12-year-old orphaned by AIDS covering topics about AIDS-related loss, HIV transmission and treatment, disclosure, youth identity, adherence, stigma, communication, puberty, safety strategies, and social support | Group and individual sessions | Every other week for 3 months | Trained lay counselors | Family-based and support | With caregivers | CHAMP model |
| Naar-King et al (2010) [17] | Healthy Choices: 4 60-minute sessions focused on helping youth address two of three possible problem behaviors based on their entry screening: substance use, sexual risk, or medication adherence | Individual sessions | Weekly for 4 weeks | Doctoral psychology students or trained mental health clinicians | Psychotherapy and behavioral | Without caregivers | Motivational Enhancement Therapy (MET) |
| Lyon et al (2010) [18] | Family-Centered Advance Care (FACE): 3 weekly 60-90-minute semi-structured family interview sessions including Lyon Advance Care Planning Survey-Adolescent and Surrogate Versions, The Respecting Choices Interview, and Completion of The Five Wishes | Group sessions | Weekly for 3 weeks | Trained or certified interviewer | Family-based and support | With caregivers | Integration of evidence-based approaches presented in the literature |

### Efficacy of the included interventions (Figure 2)

**Figure 2:**
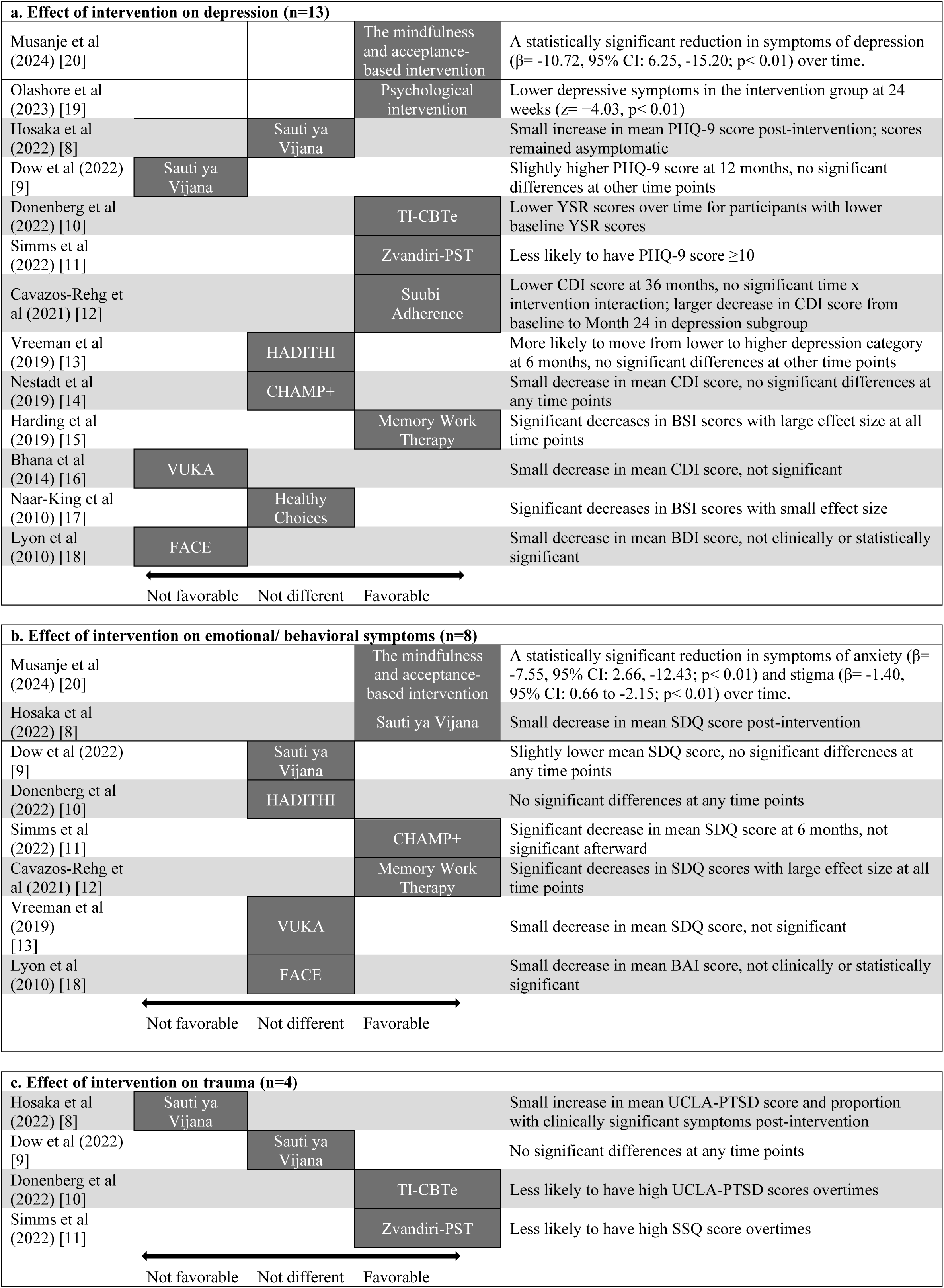
Efficacy of the included interventions.

The efficacy of the various mental health interventions for adolescents living with HIV showed mixed results across different outcome measures. For depression, interventions that demonstrated significant improvements in depressive symptoms were mindfulness and acceptance-based intervention, psychoeducation intervention, Memory Work Therapy, Zvandiri Problem-solving therapy (Zvandiri-PST), and Trauma-Informed Cognitive Behavioral Therapy (TI-CBTe), while Sauti ya Vijana and Helping AMPATH Disclose Information and Talk about HIV Infection (HADITHI) showed limited or mixed efficacy. For emotional and behavioral symptoms, the mindfulness and acceptance-based intervention, Memory Work Therapy showed the most consistent positive impact, with significant decreases in The Strengths and Difficulties Questionnaire (SDQ) scores and large effect sizes. For trauma-related outcomes, the results were less consistent, with TI-CBTe and Zvandiri-PST showing some positive impact on the University of California Los Angeles post-traumatic stress disorder (UCLA-PTSD) and Shona Symptom Questionnaire (SSQ) scores, respectively.

## Discussion

This review included 13 studies with diverse participant demographics, highlighting the global importance of addressing mental health in adolescents living with HIV. The interventions identified were varied and demonstrated effectiveness, although the magnitude and duration of their effects were inconsistent. The studies collectively underscored the necessity of targeting various mental health aspects, such as depression, anxiety, trauma, and behavioral symptoms, despite employing different scales to measure outcomes. The varied follow-up periods and analysis methods, however, complicated direct comparisons of intervention effectiveness, illustrating the complexity of mental health care for young people living with HIV.

Several factors potentially contributed to the mixed results observed in this review. First, the interventions themselves varied in their theoretical foundations, components, and delivery methods, which might affect their effectiveness. Second, the included studies were diverse in terms of sample size, study design, and outcome measures, complicating definitive conclusions about intervention effectiveness. Third, contextual factors such as cultural, social, and economic factors, along with differences in HIV care settings, might influence the generalizability and applicability of the interventions across various populations and contexts. Fourth, for most studies, psychiatric symptoms were not elevated at baseline, making it harder to show a clear benefit. Similar results patterns were observed in related literature reviews [4, 13].

Literature gaps identified in this review include the limited number of studies conducted in certain regions, such as Latin America, Europe, or Asia, as well as the lack of research on specific subpopulations, like adolescents at different HIV infection stages or those with specific comorbid mental health conditions. Moreover, the long-term effectiveness of these interventions was inadequately assessed, and few studies compared the effectiveness of different intervention components or delivery formats.

Despite these limitations and gaps, this review offered valuable insights into the current state of research on mental health interventions for adolescents living with HIV. The review identified characteristics of effective interventions for this population, informing the tailoring and adaptation of interventions to their unique needs, considering their developmental stage, social context, and challenges of managing HIV infection. Engaging local stakeholders, including HIV-infected adolescents, their families, and healthcare providers, may enhance the applicability of these interventions in different cultural, social, and healthcare settings, ultimately increasing their reach and impact. This scoping review built upon previous reviews by including recent studies, examining a broader range of interventions, and contemplating the potential for adapting interventions to various contexts. By synthesizing the latest evidence, this review provides a more comprehensive understanding of the state of research on mental health interventions for adolescents living with HIV, offering an evidence-informed guide for future research and practice.

## Conclusion

Evidence-based interventions targeting mental health outcomes in HIV-infected adolescents have shown some effectiveness, but further research is needed to determine their long-term impact and the best strategies for implementation. This scoping review provides a useful resource for addressing the mental health needs of HIV-infected adolescents and informing the selection of appropriate interventions for adaptation in other countries. By offering important insights into improving the mental health of HIV-infected adolescents, this review furnishes an evidence-informed guide for future research and practice in this area.

## Data Availability

All data produced in the present work are contained in the manuscript

## Acknowledgment

Thank you to all participants and authors in the included studies.

## Ethics approval

This review protocol has been registered with PROSPERO under registration ID CRD42022382740.

## Authors’ contribution

All members developed a protocol and reviewed and feedbacked PROSPERO registration, results, and manuscripts. Van Thi Ngoc Tran and Lam Khanh Phung searched and screened potential articles in December 2023. Van Thi Ngoc Tran and Hoa H Nguyen screened the updated articles in September 2024. Hoa H Nguyen revised manuscripts and submitted them to the journal.

